# Blood biomarker score identifies individuals at high risk for severe COVID-19 a decade prior to diagnosis: metabolic profiling of 105,000 adults in the UK Biobank

**DOI:** 10.1101/2020.07.02.20143685

**Authors:** Nightingale Health UK Biobank Initiative, Heli Julkunen, Anna Cichońska, P. Eline Slagboom, Peter Würtz

## Abstract

**Background:** Identification of healthy people at high risk for severe COVID-19 is a global health priority. We investigated whether blood biomarkers measured by high-throughput metabolomics could be predictive of severe pneumonia and COVID-19 hospitalisation years after the blood sampling.

**Methods:** Nuclear magnetic resonance metabolomics was used to quantify a comprehensive biomarker profile in 105 146 plasma samples collected in the UK Biobank during 2007–2010 (age range 39–70). The biomarkers were tested for association with severe pneumonia (2507 cases, defined as diagnosis in hospital or death record occurring during a median of 8.1-year follow-up) and with severe COVID-19 (195 cases, defined as diagnosis in hospital between mid-March to mid-June 2020). A multi-biomarker score was derived for prediction of severe pneumonia based on half of the study population and validated in the other half. We explored how this biomarker score relates to the risk of severe COVID-19.

**Findings:** The biomarker associations with risk of severe COVID-19 followed an overall pattern similar to associations with risk of severe pneumonia (correlation 0.83). The multi-biomarker score, comprised of 25 blood biomarkers including inflammatory proteins, fatty acids, amino acids and advanced lipid measures, was strongly associated with risk of severe pneumonia (odds ratio 1.67 per SD [95% confidence interval 1.59–1.76]; 3.8-fold risk increase for individuals in upper vs lower quintile). The multi-biomarker score was also associated with risk of severe COVID-19 (odds ratio 1.33 per SD [1.17–1.53]; 2.5-fold risk for upper vs lower quintile) and remained significant when adjusting for body mass index, smoking, and existing respiratory and cardiometabolic diseases. Mimicking the decade lag from blood sampling to COVID-19, severe pneumonia events occurring after 7–11 years associated with the multi-biomarker score to a similar magnitude (odds ratio 1.43 per SD [1.29–1.59]; 2.6-fold risk for upper vs lower quintile) as for severe COVID-19. Interpolating to a screening scenario today, the magnitude of association of the multi-biomarker score was 3 times higher for short-term risk of severe pneumonia (odds ratio 2.21 per SD [1.95–2.50]; 8.0-fold risk for upper vs lower quintile in analysis of events during first 2 years after blood sampling).

**Interpretation:** In decade-old blood samples from the UK Biobank, a multi-biomarker score measured by high-throughput metabolomics is indicative of the risk for severe COVID-19. The molecular signature of biomarker changes reflective of risk for severe COVID-19 is similar to that for severe pneumonia, in particular when accounting for the time lag to the COVID-19 pandemic. The even stronger association of the biomarker score with 2-year risk for severe pneumonia lends support to promising screening possibilities for identifying people at high risk for severe COVID-19.

## Introduction

The coronavirus disease 2019 (COVID-19) pandemic represents an unprecedented threat globally. Protection of individuals at the highest risk for severe and potentially fatal COVID-19 is a prime component of national policies, with stricter social distancing and other preventative means currently recommended mainly for elderly people and individuals with chronic health conditions. However, seemingly healthy middle-aged individuals also suffer from severe COVID-19 (Zhou et al 2020, Atkins et al 2020, OpenSAFELY Collaborative 2020). More accurate identification of apparently healthy individuals at high risk could help to target social distancing as lockdown measures are being relaxed, facilitate safe return to work, and eventually prioritise individuals for vaccines with limited supply.

Pneumonia is a life-threatening complication of COVID-19 and the most common diagnosis in severe COVID-19 patients. As for COVID-19, the main risk factors for community-acquired pneumonia are high age and pre-existing respiratory and cardiometabolic diseases that can weaken the lungs and the immune system (Almirall et al 2017). Based on analyses of large blood sample collections of healthy individuals, the biomarkers predictive of severe COVID-19 are partly shared with biomarkers reflective of the risk for contracting pneumonia, including lower HDL cholesterol and elevated markers of impaired kidney function and inflammation (Ho et al 2020).

Comprehensive profiling of metabolic biomarkers, also known as metabolomics, in prospective population studies have suggested a range of blood biomarkers for cardiovascular disease and diabetes to also be reflective of future risk for contracting severe infectious diseases (Ritchie et al 2015, Deelen et al 2019). Metabolic biomarker profiling of severe COVID-19 patients could therefore potentially aid high-risk identification among healthy individuals, but such studies require measurement of large numbers of pre-pandemic samples to obtain sufficient number of individuals who eventually contract COVID-19 and require hospitalisation. Metabolic biomarker profiles have recently been measured using Nightingale nuclear magnetic resonance (NMR) metabolomics in over 100 000 plasma samples from the UK Biobank, with clinical data linkage unlocked 22 May 2020.

Here, we examined if metabolic biomarkers measured from blood samples collected a decade before the pandemic could be predictive of COVID-19 hospitalisation in UK general population settings. Exploiting the shared risk factor relation between COVID-19 and pneumonia (Hu et al 2020), we used well-powered statistical analyses of biomarkers with severe pneumonia risk to develop a multi-biomarker score that condenses the information from the metabolic profile into a single metric. Taking advantage of the time-resolved information on the occurrence of severe pneumonia events in the UK Biobank, we mimicked the influence of the decade lag from blood sampling to the COVID-19 pandemic on the biomarker associations, and used short-term follow-up to interpolate to a scenario of preventative COVID-19 screening carried out today.

## Methods

### Study population

Details of the design of the UK Biobank have been reported previously (Sudlow et al 2015). Briefly, the UK Biobank recruited 502 639 participants aged 37–70 years in 22 assessment centres across the UK. All participants provided written informed consent and ethical approval was obtained from the North West Multi-Center Research Ethics Committee. Blood samples were drawn at baseline between 2007 and 2010. The current analysis was approved under UK Biobank Project 30418. No selection criteria were applied to the sampling.

### Metabolic biomarker profiling

From the entire UK Biobank population, a random subset of non-fasting baseline plasma samples (aliquot 3) from 118 466 individuals and 1298 repeat-visit samples were measured using targeted high-throughput NMR metabolomics (Nightingale Health Ltd; biomarker quantification version 2020). This provides simultaneous quantification of 249 metabolic measures, including routine lipids, lipoprotein subclass profiling with lipid concentrations within 14 subclasses, fatty acid composition, and various low-molecular weight metabolites such as amino acids, ketone bodies and glycolysis metabolites quantified in molar concentration units. Technical details and epidemiological applications have been reviewed (Soininen et al 2015, Würtz et al 2017). The Nightingale NMR metabolomics technology has received various regulatory approvals, including CE-mark, and 37 biomarkers in the panel have been certified for diagnostics use (marked in (**Supplementary Table 1**).. We focused on this set of clinically validated biomarkers to facilitate potential translational applications and since these biomarkers span most of the different metabolic pathways measured by the NMR metabolomics platform. The metabolomics data were curated and linked to UK Biobank clinical data in late-May 2020.

**Table 1.** Clinical characteristics of the UK Biobank participants in the current study.

|  | Severe pneumonia<br>(diagnosis in hospital<br>or death record) |  | Severe COVID-19<br>(diagnosis in hospital) |  |
| --- | --- | --- | --- | --- |
|  | Incident<br>cases | Controls | Incident<br>cases | Controls |
| Individuals with NMR biomarker<br>measures | 2507 | 102 639 | 195 | 92 452 |
| Age at blood sampling (median,<br>[range]) | 62<br>[40-70] | 58<br>[39-70] | 60<br>[40-70] | 58<br>[39-70] |
| Females (%) | 44 % | 54 % | 41 % | 54 % |
| Body mass index (mean, kg/m<sup>2</sup>) | 28.5 | 27.4 | 28.8 | 27.3 |
| Proportion with prevalent chronic respiratory and cardiometabolic diseases |  |  |  |  |
| Cardiovascular disease (%) | 17.5 % | 6.6 % | 16.9 % | 6.5 % |
| Diabetes (%) | 9.3 % | 3.9 % | 9.2 % | 3.9 % |
| Lung cancer (%) | 0.4 % | 0.1 % | 0.5 % | 0.1 % |
| Chronic obstructive pulmonary<br>disease (%) | 6.1 % | 0.7 % | 2.6 % | 0.8 % |
| Liver diseases (%) | 1.5 % | 0.7 % | 2.1 % | 0.7 % |
| Renal failure (%) | 3.6 % | 1.3 % | 3.6 % | 1.4 % |

### Severe pneumonia outcomes

We combined ICD-10 codes J12–J18 to define the pneumonia endpoint. To strengthen the analogy with analysis of severe COVID-19, we focused on severe pneumonia events, defined as diagnosis in hospital or death record based on UK Hospital Episode Statistics data and national death registries (2507 incident cases in the current study). All analyses are based on first occurrence of diagnosis, and therefore 2658 individuals with recorded hospitalisation of pneumonia prior to blood sampling were excluded. Additionally, 346 individuals with pneumonia diagnoses recorded in primary care settings and by self-reports were also omitted from the analyses. The registry-based follow-up was from blood sampling in 2007–2010 through to 2016–2017, depending on assessment centre (850 000 person-years). Due to the shorter follow-up for severe pneumonia, there were no overlapping cases with severe COVID-19.

### Severe COVID-19 outcomes

We used COVID-19 data available in the UK Biobank per 22 June 2020, which covers test results from 16 March to 14 June 2020. These data include information on positive/negative PCR-based diagnosis results and explicit evidence in the microbiological record on whether the participant was an inpatient (UK Biobank Data Resource, 2020). For the present analyses, we focused on PCR-positive inpatient diagnoses. These hospitalised cases are here denoted as severe COVID-19 (195 cases in the current study). There were additionally 80 test-positive individuals without explicit evidence of being an inpatient, which were inferred as non-severe and omitted from present analyses. COVID-19 data were not available for assessment centres in Scotland and Wales, so individuals from these centres were excluded. Individuals who had died during follow-up prior to 2018 were also excluded, since they were never exposed to COVID-19.

### Prevalent respiratory and cardiometabolic disease

To examine the influence of prevalent diseases in the prospective analyses of severe pneumonia and severe COVID-19, we used the following definitions of chronic respiratory and cardiometabolic conditions: prevalent cardiovascular disease (ICD-10 codes I20–I25, I50, I60–I64 and G45), diabetes (E10–E14), lung cancer (C33–C34, D02.2, Z85.1), chronic obstructive pulmonary disease (COPD; J43–J44), liver diseases (K70–K77), and renal failure (N17–N19).

### Statistical methods

Biomarker levels outside four interquartile ranges from median were considered as outliers and excluded. All 37 biomarkers were scaled to standard deviation (SD) units prior to analyses. For biomarker association testing with severe pneumonia and with severe COVID-19 (as separate outcomes), we used logistic regression models adjusted for age, sex, and assessment centre. To examine the utility of multiple biomarkers in combination, we used a weighted sum of the biomarkers optimised for prediction of severe pneumonia; this multi-biomarker score was denoted as ‘infectious disease score’. To minimize the collinearity of the biomarkers, the multi-biomarker score was trained using logistic regression with least absolute shrinkage and selection operator (LASSO), which uses L1 regularization that adds penalty equal to the absolute value of the magnitude of the coefficients. The multi-biomarker infectious disease score was trained using half of the study population with complete data available for the 37 clinically validated biomarkers (n=52 573 and 1257 severe pneumonia events) using five-fold cross-validation to optimize the regularization parameter *λ*. The remaining half of the study population was used in validating the performance of the biomarker score in relation to risk for severe pneumonia. The multi-biomarker infectious disease score was then tested for association with severe pneumonia and COVID-19 in logistic regression models adjusted for age, sex, and assessment centre. We further examined additional adjustment for body mass index (BMI) and smoking status (never, former, current), as well as prevalent respiratory and cardiometabolic diseases. The associations were also examined by omitting individuals with prevalent respiratory and cardiometabolic diseases and stratified by age and sex. In the case of severe pneumonia, we further examined the association magnitudes according to follow-up time: we used severe pneumonia events that occurred during 7–11 years after the blood sampling to mimic the decade long lag from blood sampling to the COVID-19 pandemic, and severe pneumonia events occurring within the first 2 years to interpolate to the scenario of preventative screening for COVID-19 carried out today (in both scenarios the confined follow-up times were arbitrarily chosen as short as possible while ensuring sufficient number of events). Finally, to explore potential non-linear effects, the infectious disease score was plotted as a proportion of individuals who contracted severe pneumonia during follow-up when binning individuals into percentiles of the infectious disease score (Khera et al 2018) and the time-resolution was examined by Kaplan-Meier curves of the cumulative risk for severe pneumonia.

## Results

Characteristics of the study population are shown in **Table 1**. Among the 105 146 UK Biobank study participants with complete data on metabolic biomarkers and severe pneumonia outcomes, and no prior history of diagnosed pneumonia, there were 2507 severe pneumonia events recorded in hospital or death registries after the baseline blood sampling (median follow-up time 8.1 years).

For the severe COVID-19 analyses, there were 195 PCR-confirmed positive cases diagnosed in hospital (inferred as severe cases in this study) among the 92 647 individuals with COVID-19 data linkage available per 22 June 2020. In June 2020, the age range of study participants was 49–84 years. The median duration from blood sampling to the COVID-19 pandemic was 11.2 years (interquartile range 10.0–12.6). The prevalence of chronic respiratory and cardiometabolic diseases was similar for study participants who developed severe pneumonia and those who contracted COVID-19 and required hospitalisation; there was no overlap in cases.

The number of individuals analysed for severe COVID-19 is slightly lower than for severe pneumonia, since no COVID-19 data were available from assessment centres in Scotland and Wales.

### Metabolic biomarkers and severe pneumonia risk

**Figure 1A** shows the associations of 37 biomarkers with risk of severe pneumonia in the entire study population (n=105 146). The biomarkers highlighted here are those with regulatory approval for diagnostics use in the Nightingale NMR metabolomics platform; results for all 249 metabolic measures quantified are shown in **Supplementary Figure 1**. Strong associations were observed across several metabolic pathways: increased plasma concentrations of cholesterol measures, omega-3 and omega-6 fatty acid levels, histidine, branched-chain amino acids and albumin were associated with lower risk for contracting severe pneumonia. Increased concentrations of mono-unsaturated and saturated fatty acids, as well glycoprotein acetyls (GlycA, a marker of chronic inflammation) were associated with elevated risk for contracting severe pneumonia.

**Figure 1.**
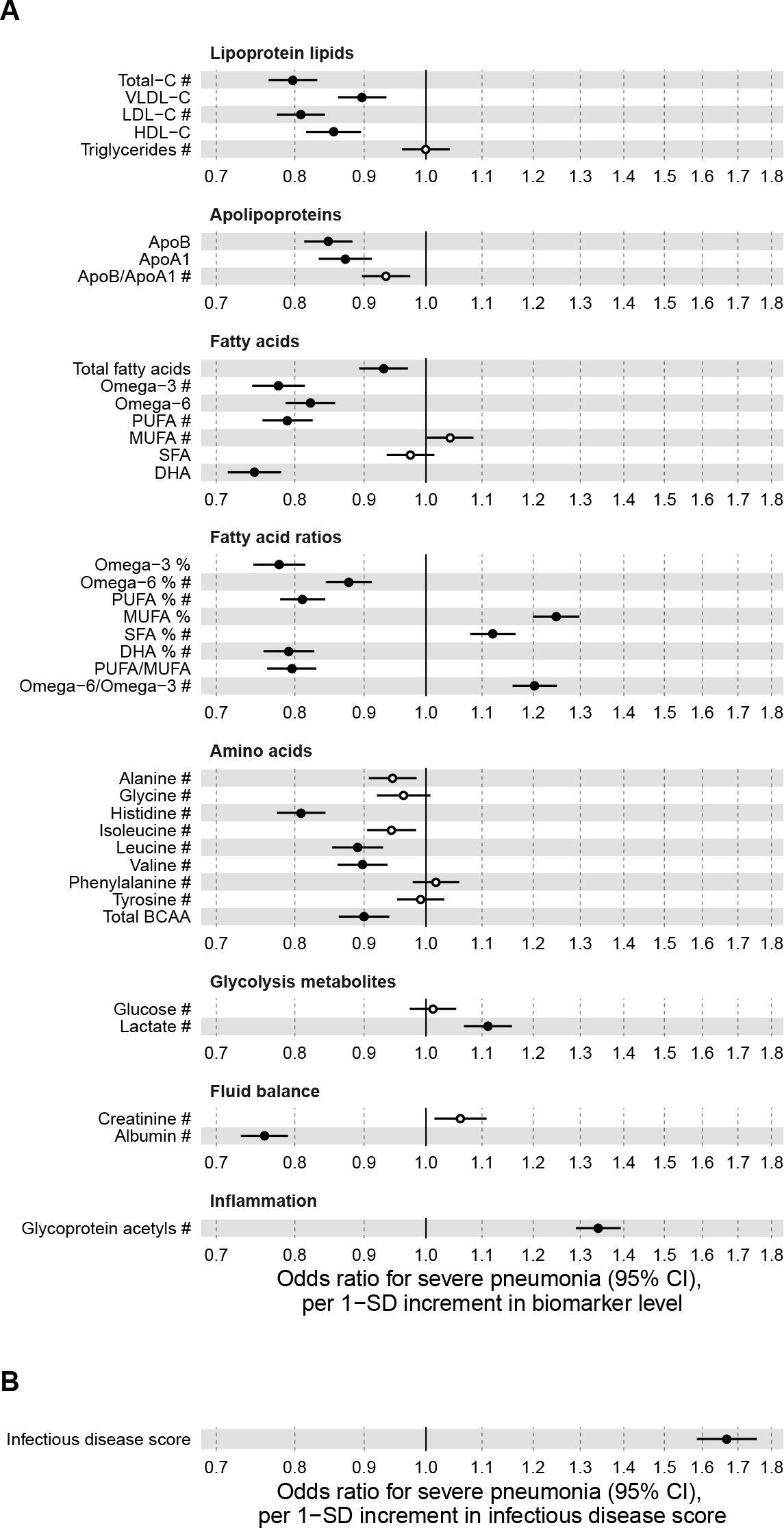
Relation of baseline biomarker and multi-biomarker infectious disease score to risk of severe pneumonia (n=105 146; 2507 incident events). (A) Odds ratios with severe pneumonia (2507 hospitalisations or deaths during a median of 8 years of follow-up) for 37 clinically validated biomarkers measured by Nightingale NMR metabolomics. (B) Odds ratio with severe pneumonia for the multi-biomarker infectious disease score. The infectious disease score is the weighted sum of 25 out of 37 clinically validated biomarkers, optimised for prediction of severe pneumonia based on one half of the study population using LASSO regression. Biomarkers included in the infectious disease score are marked by #. The odds ratio for infectious disease score is evaluated in the other half of the study population (n=52 573; 1250 events). All models are adjusted for age, sex, and assessment centre. Odds ratios are per 1-SD increment in the biomarker levels. Horizontal bars denote 95% confidence intervals. Closed circles denote P-value < 0.001 and open circles P-value 0.001. BCAA indicates branched-chain amino acids; DHA: docosahexaenoic acid; MUFA: monounsaturated fatty acids; PUFA: polyunsaturated fatty acids; SFA: saturated fatty acids.

Since all the biomarkers are quantified in the same single measurement, we examined if even stronger associations with severe pneumonia could be obtained using a multi-biomarker combination. We derived this multi-biomarker combination, denoted ‘infectious disease score’, using logistic regression with LASSO for variable selection, considering the 37 clinically validated biomarkers in half of the study population. This resulted in an infectious disease score comprised of the weighted sum of 25 biomarkers, with the weights selected by the machine learning algorithm. The strongest weight was attributed to GlycA (**Supplementary Table 2**).

The multi-biomarker infectious disease score was then tested for association with severe pneumonia in the other half of the study population. The magnitude of association for the infectious disease score was approximately twice as strong with severe pneumonia compared to any of the individual biomarkers (**Figure 1B**). The odds for contracting severe pneumonia was increased 67% per 1-SD increment in the infectious disease score. This corresponds to close to 4-fold higher risk for severe pneumonia among people in the upper quintile of the infectious disease score, compared to those with a score in the lowest quintile.

To assess the robustness of association with severe pneumonia, we adjusted for BMI, smoking, and existing diseases, and performed analyses stratified by age and sex (**Figure 2**). The association was attenuated by ∼10% in magnitude when adjusting for, or omitting, individuals with a diagnosis of respiratory and cardiometabolic disease at time of blood sampling (cardiovascular disease, diabetes, lung cancer, COPD, liver diseases, and renal failure; panel 2A and 2B). The association was similar across age groups, and also for men and women analysed separately (panels 2C and 2D).

**Figure 2.**
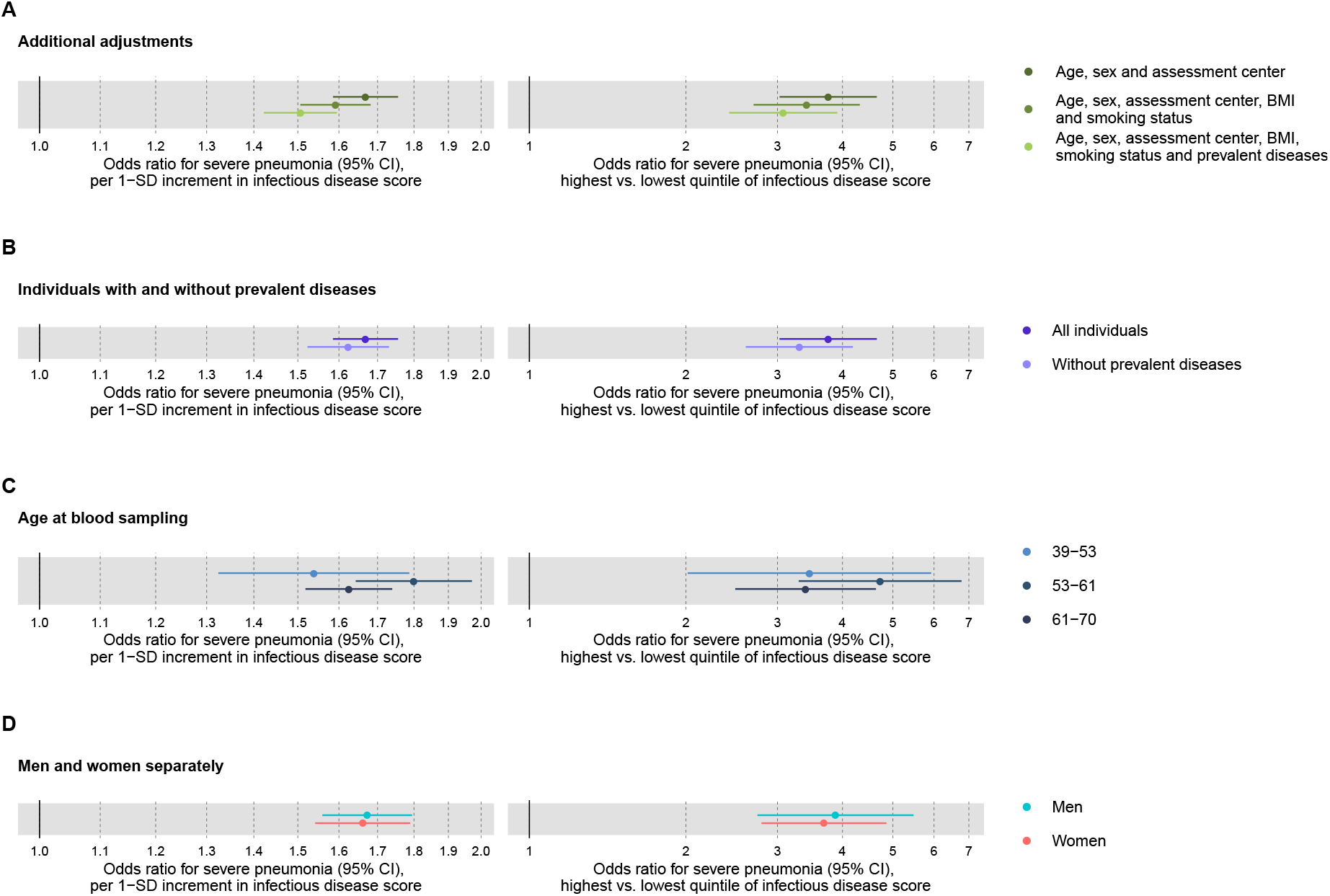
Relation of the multi-biomarker infectious disease score to risk of severe pneumonia with additional adjustments and in subgroups (n=52 573; 1250 incident events). (A) Odds ratios with severe pneumonia after additional adjustment for BMI, smoking status, and prevalent respiratory and cardiometabolic diseases. (B) Odds ratios with severe pneumonia in study participants with and without prevalent respiratory and cardiometabolic disease. (C) Odds ratios by age tertiles at the time of blood sampling. (D) Odds ratios for men and women separately. All models are adjusted for age, sex, and assessment centre. The left-hand side shows odds ratio per 1-SD increment in the multi-biomarker infectious disease score, and the right-hand side odds ratios for individuals in the upper vs lower quintiles of the score. The results are based on the validation-half of the study population that was not used to derive the infectious disease score (1250 events during a median of 8 years of follow-up).

To mimic the influence of the decade-long lag from blood sample collection to the COVID-19 pandemic, we tested the association of the infectious disease score with severe pneumonia events occurring during 7–11 years after the blood sampling (**Figure 3A**). Since there were only few severe pneumonia events recorded with more than 9 years of follow-up, we could not fully mimic the decade long time lag to the COVID-19 pandemic. The association magnitude for this time-lag accounting scenario was almost half of that observed for events occurring within the first 7 years. To interpolate to a scenario of preventative screening conducted today, we also tested the short-term association with severe pneumonia events occurring within the first 2 years after the blood sampling (**Figure 3B**). The association magnitude in this scenario was approximately twice as strong as for severe pneumonia events occurring at least 2 years after blood sampling (odds ratio 2.2 per 1 SD; 8.0-fold risk elevation for individuals in the upper vs lower quintile of the infectious disease score). The elevation in short-term risk for severe pneumonia for high levels of the infectious disease score remained strong when adjusting for BMI, smoking and prevalent respiratory and cardiometabolic diseases (6.1-fold risk elevation for individuals in the upper vs lower quintile; **Supplementary Figure 2**).

**Figure 3.**
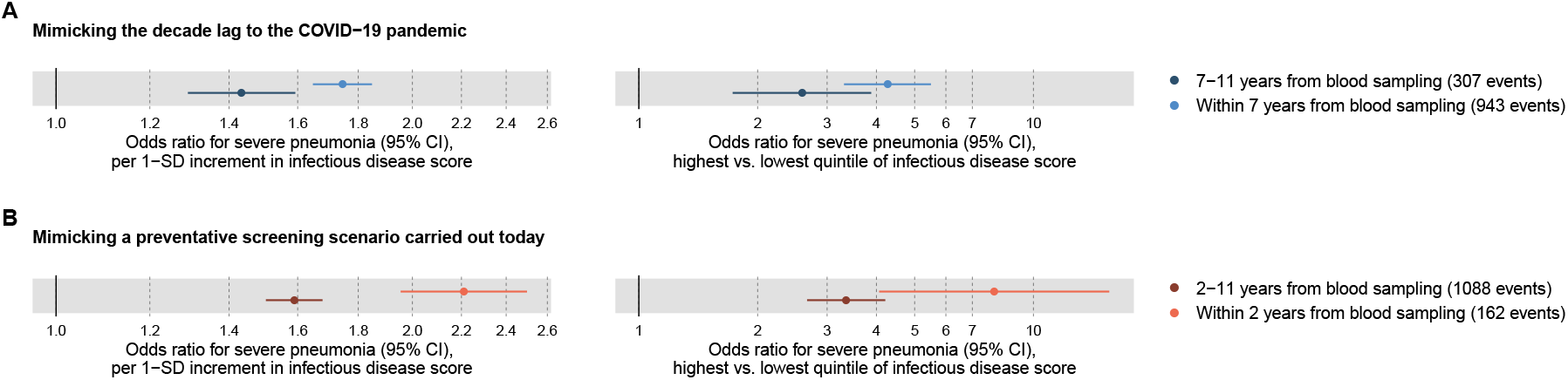
Relation of the multi-biomarker infectious disease score to long-term and short-term risk for severe pneumonia (n=52 573; 1250 incident events). (A) Odds ratios with severe pneumonia events occurring within the first 7 years of blood sampling, compared to events that occurred 7-11 years after blood sampling. (B) Odds ratios for severe pneumonia occurring within and after the first 2 years of blood sampling. Models are adjusted for age, sex, and assessment centre. The left-hand side shows odds ratio per 1-SD increment in the multi-biomarker infectious disease score, and the right-hand side odds ratios for individuals in the upper vs lower quintiles of the score. The results are based on the validation-half of the study population that was not used to derive the infectious disease score

We next explored the risk gradient for severe pneumonia along increasing levels of the infectious disease score, since non-linear effects could potentially facilitate identification of thresholds for high risk. **Figure 4A** shows the increase in proportion of individuals who contracted severe pneumonia according to percentiles of the score. The risk increased prominently in the upper quintile, and particularly for the highest few percentiles. The time-resolved plot of the cumulative probability of severe pneumonia during follow-up is shown in **Figure 4B**. The increase in risk for severe pneumonia was elevated among individuals with the very highest levels of multi-biomarker infectious disease scores, and this was observed already during the first few years of follow-up, corroborating the time-censored odds ratios given in Figure 3. The prominent and immediate risk elevation was also observed when limiting analyses to individuals without chronic respiratory and cardiometabolic diseases at time of blood sampling (**Supplementary Figure 3**).

**Figure 4.**
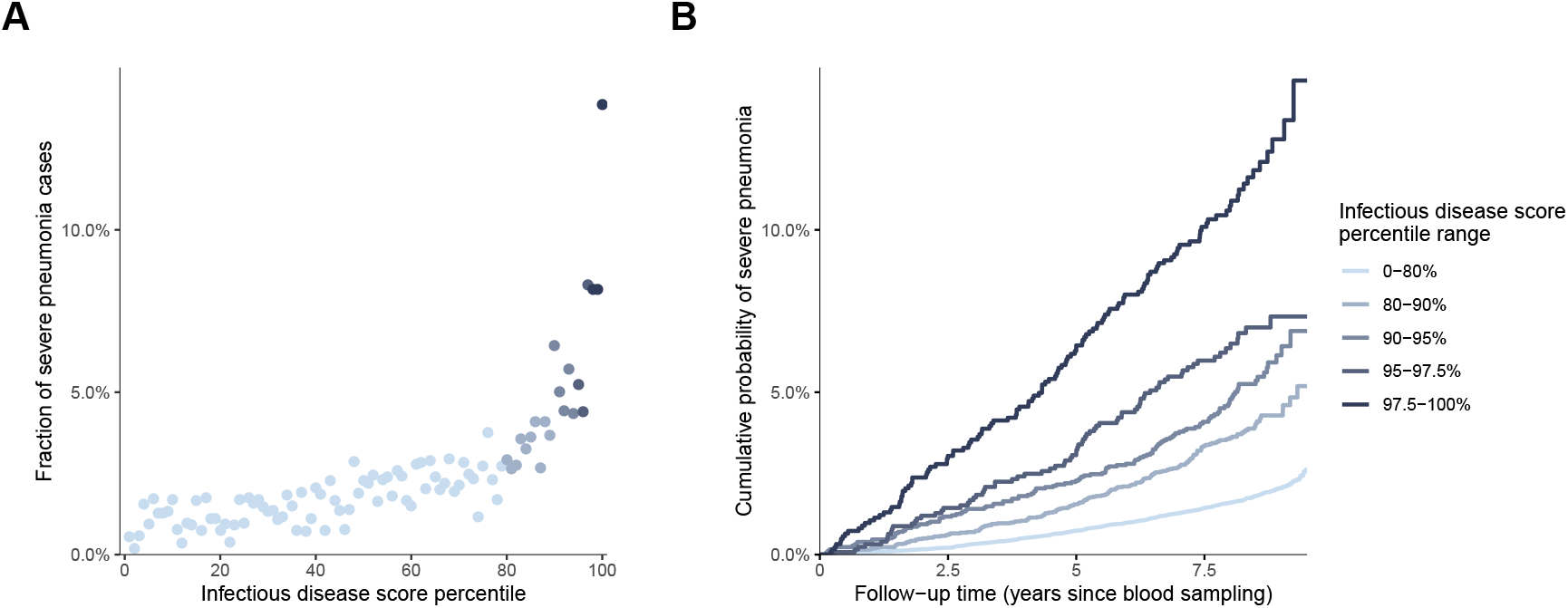
Risk gradient for contracting severe pneumonia according to percentiles of the multi-biomarker infectious disease score (n=52 573; 1250 incident events). (A) Proportion of individuals who contracted severe pneumonia during a median of 8 years of follow-up according to the percentile of the multi-biomarker infectious disease score. Each point represents approximately 500 individuals. (B) Kaplan-Meier curves of the cumulative probability of severe pneumonia in quantiles of the multi-biomarker infectious disease score. The follow-up time was truncated at 9.5 years since only a small fraction of individuals were followed longer. Results are based on the validation half of the study population that was not used for deriving the infectious disease score (n=52 573). The corresponding plots for individuals free of baseline respiratory and cardiometabolic diseases are shown in Figure S3.

### Metabolic biomarkers and severe COVID-19

**Figure 5** shows the associations of the 37 clinically validated biomarkers and the infectious disease score with severe COVID-19 (defined as PCR-confirmed positive inpatient diagnosis). Many of the individual biomarkers were associated with increased risk for severe COVID-19 at P-value<0.05, including lower levels of omega-3 and omega-6 fatty acids as well as albumin, and higher levels of GlycA. We observed a high concordance in the overall pattern of biomarker associations with severe pneumonia (cf. Figure 1A), with a Spearman correlation of 0.83 between the overall biomarker signatures for severe pneumonia and severe COVID-19 (**Figure 6**). The infectious disease score was robustly associated with severe COVID-19, with the odds increased 34% per 1-SD. This corresponds to 2.5 fold higher risk for severe COVID-19 for the individuals in the upper quintile of the infectious disease score, compared to individuals whose score was in the lowest quintile. This magnitude of association was similar to that observed with severe pneumonia events occurring during the interval of 7–11 years after the blood sampling.

**Figure 5.**
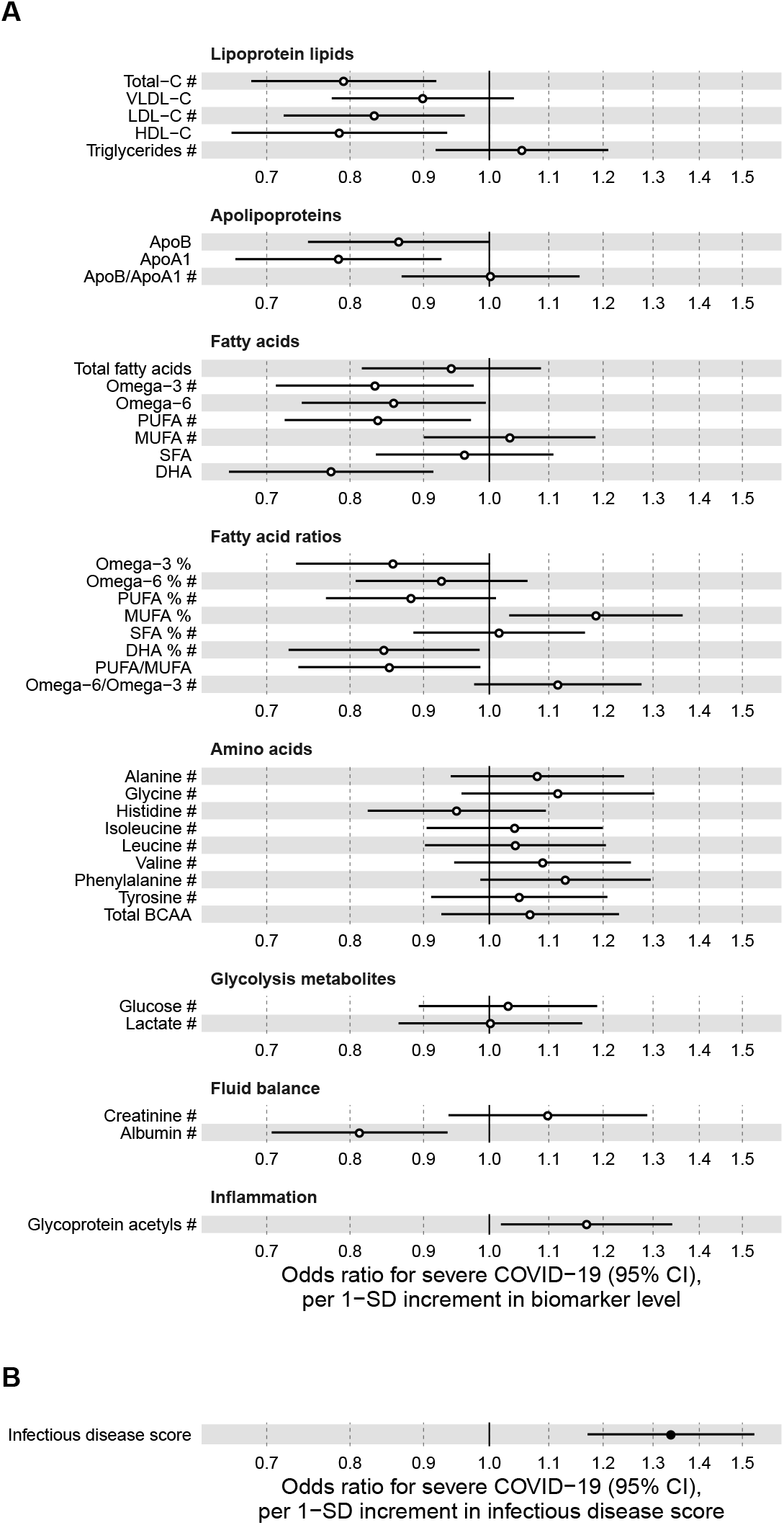
Relation of baseline biomarkers and multi-biomarker infectious disease score to risk of severe COVID-19 (n=92 647; 195 cases diagnosed in hospital). (A) Odds ratios with severe COVID-19 (defined as PCR-positivediagnosis in hospital; 195 cases out of 92 647 individuals) for 37 clinically validated biomarkers measured by NMR metabolomics. (B) Odds ratio with severe COVID-19 for the multi-biomarker infectious disease score. Biomarkers included in the infectious disease score are marked by #. Odds ratios are per 1-SD increment in the biomarker levels. Models are adjusted for age, sex, and assessment centre.

**Figure 6.**
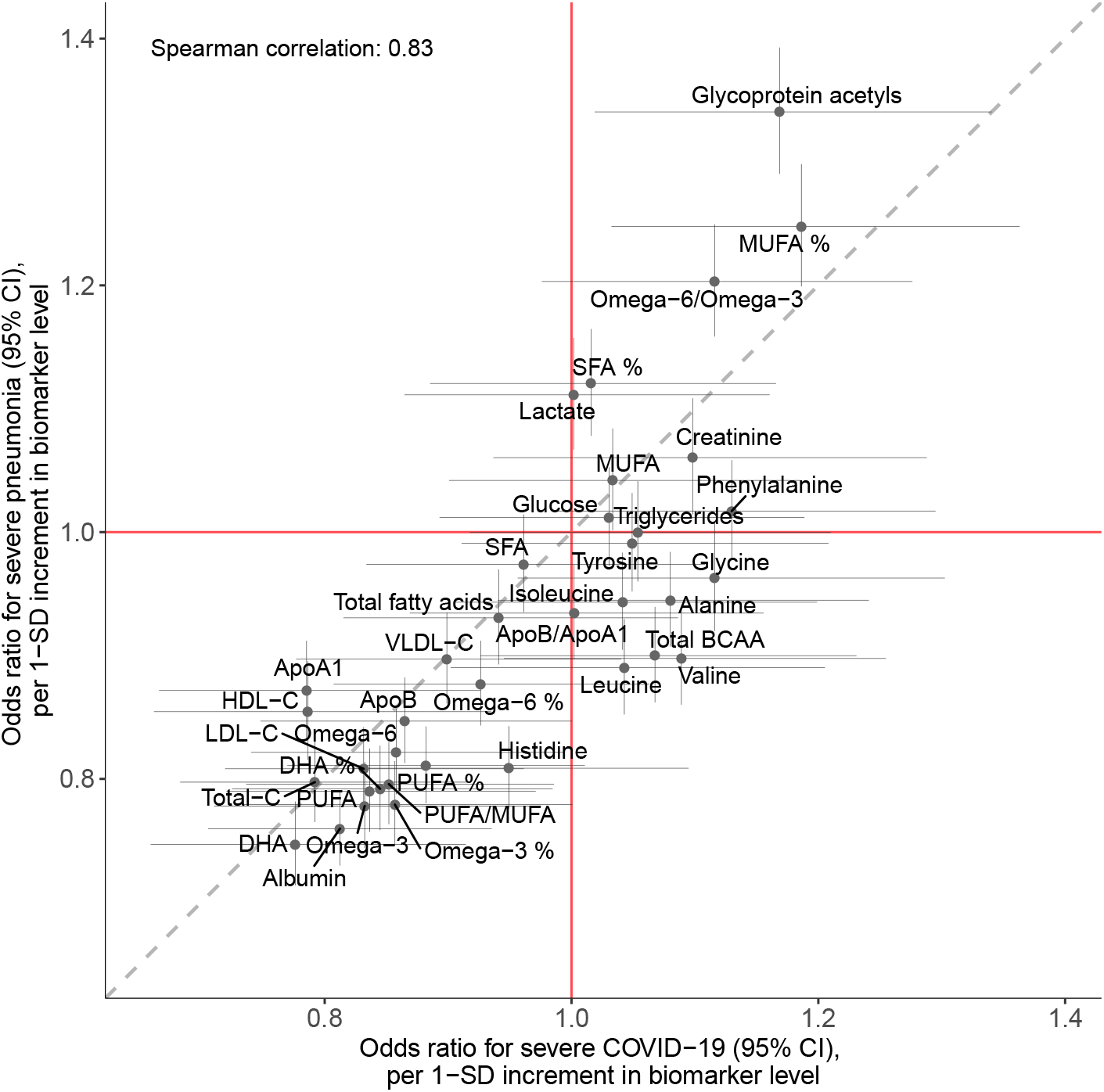
Concordance of the overall pattern of biomarker associations with severe pneumonia and severe COVID-19. Biomarker associations with severe pneumonia (y-axis) plotted against the corresponding associations with severe COVID-19 (x-axis). The odds ratio, with adjustment for age, sex, and assessment centre, for each of the 37 clinically validated biomarkers in the Nightingale NMR metabolomics panel is given with 95% confidence intervals in vertical and horizontal error bars. The dashed line denotes the diagonal.

We further examined the association of the infectious disease score with severe COVID-19 after adjustment or exclusion for prevalent disease, and conducted stratified analyses for age and sex (**Figure 7**). The association with severe COVID-19 was attenuated, but remained significant when adjusted for BMI, smoking as well as prevalent respiratory and cardiometabolic diseases (panel 7A). The associations were approximately 40% weaker when limiting the COVID-19 analyses to individuals without chronic respiratory and cardiometabolic diseases at time of blood sampling (panel 7B). There was no evidence of differences in association magnitude according to age (panel 7C) and odd ratios were similar for men and women (panel 7D).

**Figure 7.**
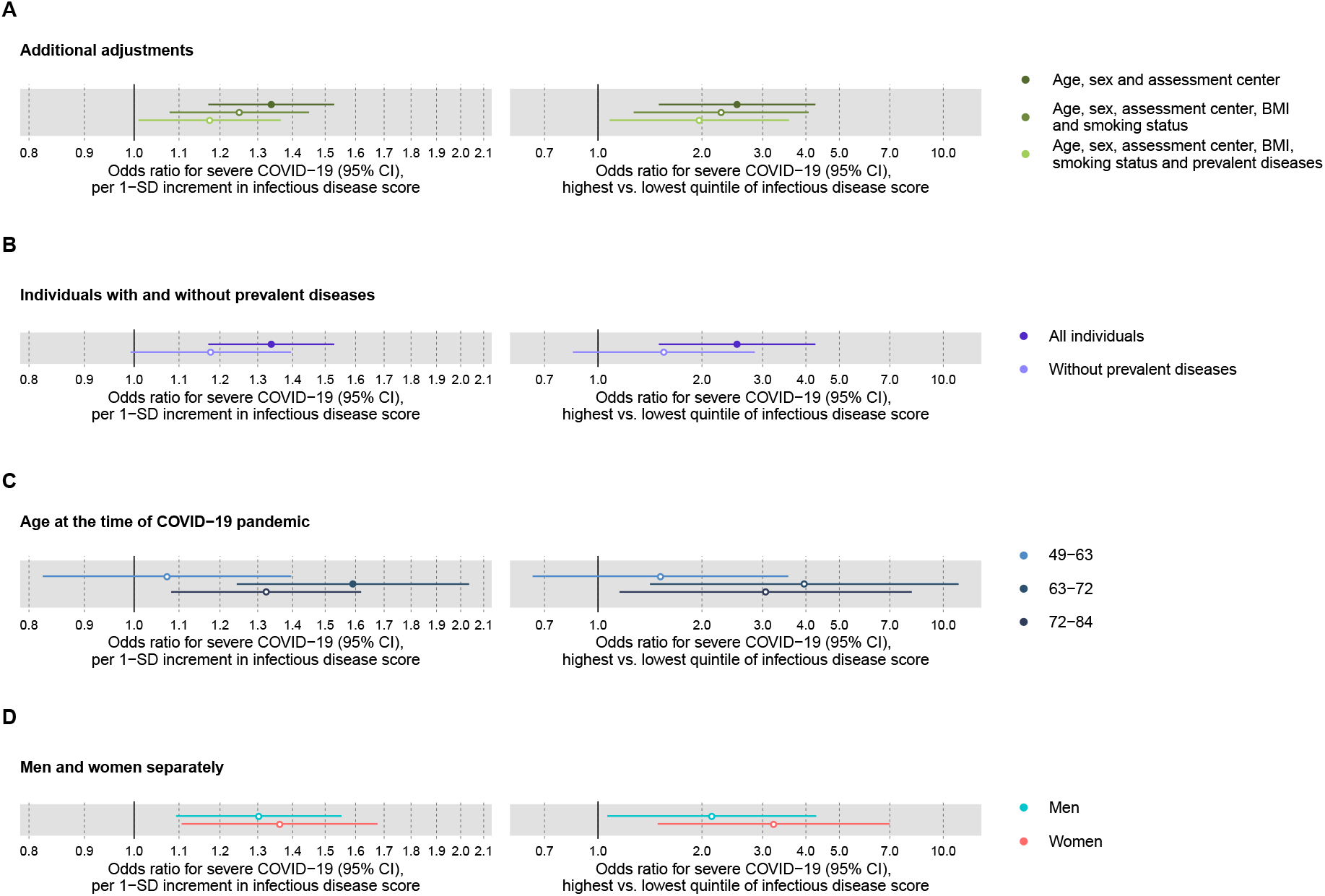
Relation of the multi-biomarker infectious disease score to risk of severe COVID-19 with additional adjustments and in subgroups of the study population (n=92 647; 195 cases diagnosed in hospital). (A) Odds ratios with severe COVID-19 after additional adjustment for BMI, smoking status, prevalent respiratory and cardiometabolic diseases. (B) Odds ratios with severe pneumonia in study participants with and without chronic cardiometabolic disease conditions at time of blood sampling. (C) Odds ratios by age tertiles at the time of the COVID-19 pandemic. (D) Odds ratios for men and women separately. Models are adjusted for age, sex and assessment centre. The left-hand side shows the odds ratio per 1-SD increment in the multi-biomarker infectious disease score, and the right-hand side the odds ratios for individuals in the upper vs lower quintiles of the score.

Finally, we examined the technical repeatability and biological stability of measuring the infectious disease score. The measurement repeatability was high (Pearson correlation 0.94 in blind duplicate samples). Even though the blood samples were primarily non-fasting, the levels of the infectious disease score were broadly stable over time in repeat visits (Pearson correlation 0.61 between baseline and repeat measurements after ∼4 years; **Supplementary Figure 4**).

## Discussion

By exploring biomarker associations with severe pneumonia risk in the largest metabolomics study to date, we generated a blood biomarker score for infectious disease risk. We demonstrated that this score, comprised of multiple metabolic biomarkers captured in a single measurement, reflects an increased risk for COVID-19 hospitalisation a decade prior to the COVID-19 pandemic. The overall signatures of biomarker associations were similar for severe COVID-19 and severe pneumonia, extending prior reports of a shared risk factor basis (Hu et al 2020). Exploiting the strong statistical power and time-resolved information for analyses of severe pneumonia, we showed that the very highest levels of the multi-biomarker infectious disease score are strongly indicative of risk for severe pneumonia. The risk elevation observed in relation to severe pneumonia events occurring after 7–11 years closely matched that of severe COVID-19, where all events happened a decade after blood sampling. Yet, a preventive screening for COVID-19 risk would require more short-term prediction; when we confined analyses to severe pneumonia events occuring during the first 2 years, the risk elevation for individuals with high levels of the multi-biomarker infectious disease score was 8 fold compared to people with low levels. That is 3 times stronger than that observed for long-term risk of severe pneumonia. By analogy, if a similar enhancement in short-term risk extend to severe COVID-19, then our results indicate potential applications for more personalised COVID-19 prevention efforts, in particular among middle-aged individuals without prevalent respiratory or cardiometabolic disease. Such a predictive tool would considerably extend our detection of COVID-19 risk beyond the current notion for older persons and those with adverse health conditions.

We observed multiple blood biomarkers, commonly linked with risk for cardiovascular disease and diabetes (Soininen et al 2015, Würtz et al 2017, Holmes et al 2018, Ahola-Olli et al 2019), to be predictive of the risk for both severe pneumonia and severe COVID-19. The overall pattern of biomarker changes followed a characteristic molecular signature reflective of increased risk for severe infectious disease. The biomarkers span multiple metabolic pathways, including low lipoprotein lipids, impaired fatty acid balance, decreased amino acid levels and high inflammation. Several of these biomarkers have also been related to frailty and risk for all-cause mortality in large prospective cohort studies (Ritchie et al 2015, Deelen et al 2019). The biological mechanisms explaining why these biomarkers reflect future risk for both chronic and infectious diseases are poorly understood, but we note that the observational nature of our study does not inform whether the biomarkers are causally contributing to increase the risk, or if they are merely indirect risk markers.

To capture the predictive information of the biomarker profile in a single metric, we used machine learning to derive an ‘infectious disease score’ composed of the weighted sum of 25 biomarkers. The strongest weight was attributed to the inflammatory marker GlycA, but we caution against interpreting the relative importance from the weights, due to high correlation between the biomarkers. The shared pattern of biomarker associations for severe pneumonia and severe COVID-19, and the coherent results for the multi-biomarker infectious disease score with long-term risk, may suggest that the overall molecular signature of biomarker changes reflects an inherent susceptibility to a more severe infectious disease course (Bonafe et al 2020). The biomarker associations with risk for severe pneumonia were largely independent of prevalent disease. In particular, the prominent elevation of severe pneumonia risk in the upper tail of the multi-biomarker infectious disease score was observed even when limiting analyses to individuals without many chronic respiratory and cardiometabolic diseases. The corresponding association with severe COVID-19 was more attenuated, but wide confidence intervals make it difficult to infer whether prevalent diseases could potentially confound the multi-biomarker infectious disease score to a larger extent than for severe pneumonia. However, studies on the entire UK Biobank sample suggest that most prevalent diseases are equally strongly associated with long-term risk for pneumonia as with severe COVID-19 (Ho et al 2020).

Most biomarker studies and omics analyses on COVID–19 have focused on characterising the disease pathophysiology and potentially aid prediction of whether already infected patients progress to severe complications (Kermali et al 2020, Shen et al 2020, Messner et al 2020). A novelty of the present study is the use of metabolic profiling of pre-COVID-19 blood samples that allowed us to explore the potential for identification of high-risk individuals. Yet, even with 105 000 samples analysed and COVID-19 case numbers consistent with per capita prevalence in the UK, we had modest statistical power for the individual biomarker associations with severe COVID-19, and to explore nonlinear effects. To overcome this limitation, we took advantage of the well-powered biomarker associations with severe pneumonia. We observed prominent risk elevation for severe pneumonia in the extreme tail of the multi-biomarker infectious disease score, whereas the risk elevation was modest for approximately four fifth of the study population. These features could aid establishing high-risk threshold for potential preventative screening applications if results generalise to severe COVID-19. The technical repeatability for measuring the infectious disease score was high, further supporting potential screening applications. The biological stability of the multi-biomarker score measured years apart was similar to that for cholesterol testing, reflecting the modifiable nature of the metabolic biomarkers but still indicating a stronger stability over time than many inflammatory markers.

Our study has both strengths and limitations. Strengths include the large sample size, which enabled the analysis of risk for severe COVID-19 based on pre-pandemic blood samples from general population settings. However, the UK Biobank study participants are not fully representative of the whole UK population (Fry et al 2017); although this is generally not a concern for investigating risk associations (Keyes and Westreich 2019), it does limit power to explore effects of ethnicity and old age. We used the widely employed Nightingale metabolomics platform, which enables absolute quantification of biomarker concentrations in a low-cost and scalable setup. While other metabolomics assays may give more molecular resolution, the panel of 37 biomarkers in the Nightingale metabolomics platform has been approved for diagnostic applications and is already used in individual person applications in Finland. Limitations include the decade long duration from blood sampling to the COVID 19 pandemic. While this limits inference on how well the biomarkers predict short-term risk for severe COVID-19, our analogy with long-term risk for severe pneumonia indicates that the time lag likely attenuates the biomarker associations substantially. Conversely, the markedly strong associations for short-term risk of severe pneumonia lead us to speculate that similar enhancements in association magnitudes could also hold for severe COVID-19. This should however be further tested, in particular in light of the bacterial origin of many severe pneumonia cases and the respiratory viral origin of COVID 19. Weaker biomarker associations for severe COVID-19 compared to severe pneumonia may also arise from the UK Biobank COVID-19 data being influenced by ascertainment bias in terms of differential healthcare seeking and differential testing, whereas severe pneumonia is anticipated to have nearly complete case ascertainment (Ho et al 2020). The currently available data on inpatient status of COVID-19 cases may further be ascertained as more detailed hospital records become available, and analyses can be updated to cover more granular assessment related to COVID-19 severity status.

In conclusion, a molecular signature of perturbed blood biomarker levels is associated with the risk for both severe pneumonia and COVID-19 hospitalisation, in samples collected a decade before the pandemic. A multi-biomarker score that captures the molecular signature in a single measurements is predictive of risk for severe COVID-19, even when adjusting for existing disease conditions. If the risk elevation observed in the upper tail of the multi-biomarker score extend from severe pneumonia to severe COVID-19, then it provides tantalising evidence to support screening applications for identification of seemingly healthy high-risk individuals, including targeted social distancing and eventual prioritisation for early vaccination. Even if the positive predictive value of the multi-biomarker score is modest, so that many people classified to the high-risk category will not develop severe COVID-19, the socioeconomic cost of targeting this group for personalised shielding should be balanced against the immense costs of wider lock-downs and the dire health consequences of the alternative.

## Data Availability

The data are available from UK Biobank, with the metabolomics data becoming public for the research community in early 2021.

https://www.ukbiobank.ac.uk

https://nightingalehealth.com

## Author Declarations

All relevant ethical guidelines have been followed; any necessary IRB and/or ethics committee approvals have been obtained and details of the IRB/oversight body are included in the manuscript.

## Acknowledgments

The authors are grateful to UK Biobank for access to data to undertake this study (Project # 30418).

## References

Ahola-Olli AV, Mustelin L, Kalimeri M, et al. Circulating metabolites and the risk of type 2 diabetes: a prospective study of 11,896 young adults from four Finnish cohorts. Diabetologia. 2019;62(12):2298–2309.

Almirall J, Serra-Prat M, Bolíbar I, Balasso V. Risk Factors for Community-Acquired Pneumonia in Adults: A Systematic Review of Observational Studies. Respiration. 2017;94(3):299–311.

Atkins JL, Masoli JAH, Delgado J, et al. Preexisting comorbidities predicting severe covid-19 in older adults in the UK biobank community cohort. medRxiv 2020.05.06.20092700.

Bonafè M, Prattichizzo F, Giuliani A, et al: Why older men are the most susceptible to SARS-CoV-2 complicated outcomes. Cytokine Growth Factor Rev. 2020;S1359-6101(20)30084-8.

Deelen J, Kettunen J, Fischer K, et al. A metabolic profile of all-cause mortality risk identified in an observational study of 44,168 individuals. Nat Commun. 2019;10(1):3346.

Fry A, Littlejohns TJ, Sudlow C, et al. Comparison of Sociodemographic and Health-Related Characteristics of UK Biobank Participants With Those of the General Population. Am J Epidemiol. 2017;186(9):1026–1034.

Ho FK, Celis-Morales CA, Gray SR, et al. Modifiable and non-modifiable risk factors for COVID-19: results from UK Biobank. medRxiv 2020.04.28.20083295.

Holmes MV, Millwood IY, Kartsonaki C, et al. Lipids, Lipoproteins, and Metabolites and Risk of Myocardial Infarction and Stroke. J Am Coll Cardiol. 2018;71(6):620–632.

Kermali M, Khalsa RK, Pillai K, Ismail Z, Harky A. The role of biomarkers in diagnosis of COVID-19 − A systematic review. Life Sci. 2020;254:117788.

Keyes KM, Westreich D. UK Biobank, big data, and the consequences of non-representativeness. Lancet. 2019;393(10178):1297.

Khera AV, Chaffin M, Aragam KG, et al. Genome-wide polygenic scores for common diseases identify individuals with risk equivalent to monogenic mutations. Nat Genet. 2018;50(9):1219–1224.

Messner C, Demichev V, Wendisch D, et al. Ultra-high-throughput clinical proteomics reveals classifiers of COVID-19 infection. Cell Systems 2020.

OpenSAFELY Collaborative, Williamson E, Walker AJ, et al. OpenSAFELY: factors associated with COVID-19-related hospital death in the linked electronic health records of 17 million adult NHS patients. medRxiv 2020.05.06.20092999.

Ritchie SC, Würtz P, Nath AP, et al. The Biomarker GlycA Is Associated with Chronic Inflammation and Predicts Long-Term Risk of Severe Infection. Cell Syst. 2015;1(4):293–301. doi:10.1016/j.cels.2015.09.007

Shen B, Yi X, Sun Y, et al. Proteomic and Metabolomic Characterization of COVID-19 Patient Sera. Cell. 2020;S0092-8674(20)30627-9.

Soininen P, Kangas AJ, Würtz P, Suna T, Ala-Korpela M. Quantitative serum nuclear magnetic resonance metabolomics in cardiovascular epidemiology and genetics. Circ Cardiovasc Genet; 2015;8:192–206.

Sudlow C, Gallacher J, Allen N, et al. UK biobank: an open access resource for identifying the causes of a wide range of complex diseases of middle and old age. PLoS Med. 2015;12(3):e1001779. U Biobank Data Resource; COVID-19 test results data; http://biobank.ctsu.ox.ac.uk/crystal/exinfo.cgi?src=COVID19_tests; Accessed June 1 2020.

Würtz P, Kangas AJ, Soininen P, Lawlor DA, Davey Smith G, Ala-Korpela M. Quantitative Serum Nuclear Magnetic Resonance Metabolomics in Large-Scale Epidemiology: A Primer on -Omic Technologies. Am J Epidemiol 2017;186:1084–1096.

Zhou F, Yu T, Du R, et al. Clinical course and risk factors for mortality of adult inpatients with COVID-19 in Wuhan, China: a retrospective cohort study Lancet. 2020;395(10229):1054–1062.

